# Temporal Clinical Features for 24-Hour Landmark Prediction of In-Hospital Mortality in ICU Patients With Diabetic Neuropathy: A MIMIC-IV Study

**DOI:** 10.64898/2026.08.17.26360508

**Authors:** Janet Sanjaya, Sakshie Pathak, Yong Si, Mohammadsaeed Haghi, Nausin Tabassum Kudrot, Greg Placencia, Kamiar Alaei, Maryam Pishgar

## Abstract

Diabetic neuropathy is associated with substantial systemic disease burden, but short-term mortality risk among affected intensive care unit (ICU) patients remains difficult to characterize. We evaluated whether temporal information from the first 24 hours of ICU care improves post-landmark mortality prediction beyond severity scores and static clinical summaries. Patients aged *>* 18 years with diabetic neuropathy were identified in MIMIC-IV v3.1. A 24-hour landmark was used: only patients alive and still hospitalized at 24 hours were included, and the outcome was subsequent in-hospital death. The final cohort included 1,347 patients, including 83 deaths (6.16%). Data were divided into an 80% development set and a locked 20% test set. Feature selection, hyperparameter tuning, calibration, and threshold selection were restricted to development data. Logistic regression, random forest, and XGBoost were evaluated. Random forest had the highest development cross-validated PR-AUC and was selected for interpretation. On the locked test set, random forest achieved an AUROC of 0.851 (95% CI 0.765–0.924), PR-AUC of 0.339, and Brier score of 0.051; XGBoost and logistic regression achieved AUROCs of 0.847 and 0.806. In a post hoc strictly nested analysis, adding temporal predictors increased discrimination across all three algorithms; random-forest AUROC increased from 0.815 with severity and static predictors to 0.870 with the full temporal representation. First-day temporal information therefore showed additional prognostic value, but external validation is required before clinical use.

## 1. Introduction

Diabetic neuropathy is one of the most common chronic complications of diabetes and encompasses a heterogeneous group of peripheral and autonomic nerve disorders associated with prolonged metabolic and vascular injury [1, 2, 3]. Its clinical consequences extend well beyond neuropathic pain and sensory loss. Progressive neuropathy contributes to impaired mobility, foot ulceration, recurrent infection, and lower-extremity complications that increase long-term morbidity and healthcare use [4, 5, 6]. Neuropathy also frequently occurs in patients with a substantial burden of other diabetes-related complications and chronic comorbid disease. Population-based evidence has linked diabetic neuropathy with increased all-cause and diabetes-related mortality [7]. Among patients who subsequently require intensive care, diabetic neuropathy may therefore identify a clinically complex subgroup in whom chronic disease burden and acute physiological deterioration both contribute to short-term risk.

Mortality risk in this population is unlikely to be captured by the diagnosis of diabetic neuropathy alone. Critically ill patients with long-standing diabetes may present with renal dysfunction, cardiovascular disease, infection, metabolic disturbance, or other concurrent organ dysfunction, while autonomic and vascular complications can further complicate the course of acute illness [1, 2]. Once admitted to the intensive care unit (ICU), prognosis depends not only on pre-existing disease but also on how physiological abnormalities evolve during the early phase of critical care. Identifying patients whose risk remains high after the first day of treatment may therefore require information describing both underlying severity and the trajectory of routinely measured clinical variables.

Conventional ICU risk assessment commonly relies on severity scores and early physiological measurements [8, 9, 10]. These approaches provide useful summaries of acute illness, but a single measurement or aggregate score cannot fully describe changes occurring during the first hours of critical illness. For example, the prognostic meaning of an abnormal creatinine, blood pressure, glucose concentration, or coagulation measurement may differ depending on whether it is improving, worsening, or remaining persistently abnormal. Measurement frequency itself may also contain information about clinical concern and the intensity of monitoring. Electronic health record data allow these patterns to be represented through repeated values, within-day variability, measurement counts, and changes between early and later observations.

A growing body of critical-care research has used longitudinal electronic health record data to model such time-dependent information. ICU time-series studies have shown that repeated physiological and laboratory measurements can support dynamic risk prediction and may provide information that is lost when observations are reduced to static summaries [11, 12, 13]. Related work has used early longitudinal ICU measurements for mortality prediction in other high-risk populations [14]. In patients with diabetes, temporal glucose patterns and early glycemic trajectories have also been associated with subsequent clinical outcomes [15, 16]. These findings suggest that the first ICU day can be viewed not only as a period for collecting baseline measurements, but also as a short observation window in which evolving physiology may contribute additional prognostic information.

Machine-learning methods are well suited to examining nonlinear relationships and interactions within large electronic health record datasets [17, 18]. MIMIC-IV contains detailed information on ICU physiology, laboratory testing, treatments, comorbidities, and hospital outcomes and has consequently been used extensively for clinical prediction research [19, 20]. Disease-specific studies have applied these methods to early mortality prediction in several high-risk ICU populations [21]. More directly, Huang et al. recently developed machine-learning models for mortality prediction among ICU patients with diabetic neuropathy using MIMIC-IV and reported an AUROC of 0.780 for the best-performing random forest model [22]. Their study demonstrated that routinely collected electronic health record variables can be used for mortality risk stratification in this population.

The existence of a diabetic-neuropathy-specific prediction model, however, leaves several questions unresolved. In particular, it remains unclear whether information describing changes during the first ICU day provides useful prognostic information beyond conventional severity scores and static clinical summaries. The definition of the prediction time is also important. When predictors are collected over the first 24 hours, the model is not making a prediction at ICU admission; it is estimating risk among patients who have survived and remained under observation long enough for those data to become available. Cohort eligibility, predictor collection, and outcome assessment should therefore be aligned to the same prediction time. Otherwise, information occurring before or after the intended prediction point can become mixed within the modeling task.

Landmark analysis provides a practical framework for defining this temporal relationship [23]. At a prespecified landmark, the eligible population is restricted to patients who remain at risk, predictors are constructed only from information available before that time, and subsequent outcomes are assessed after the landmark. This distinction is particularly relevant for first-day ICU prediction, where early death or discharge may otherwise be handled inconsistently. Model evaluation also requires separation between development decisions and final performance assessment. Repeated use of the same holdout data for feature selection, algorithm comparison, threshold optimization, or calibration can produce optimistic estimates of performance. Current guidance for clinical prediction models therefore emphasizes transparent definition of the prediction problem and clear separation of model development from final evaluation [24].

In this study, we developed and evaluated machine-learning models for post-landmark in-hospital mortality among ICU patients with diabetic neuropathy using MIMIC-IV. We defined a 24-hour landmark and used only information recorded during the first ICU day to predict mortality occurring subsequently during the same hospitalization. The analysis addressed two related questions: how accurately post-landmark mortality could be predicted using first-day clinical data, and whether temporal features provided additional predictive information compared with conventional severity scores and non-temporal first-day summaries. Logistic regression, random forest, and XGBoost were evaluated using a development cohort and a locked test set, with preprocessing, feature selection, hyperparameter tuning, calibration, and threshold selection restricted to the development data.

## 2. Methods

### 2.1. Study Design and Data Source

This retrospective prognostic modeling study used MIMIC-IV version 3.1, a de-identified electronic health record database containing hospital and intensive care data from Beth Israel Deaconess Medical Center [19, 20]. MIMIC-IV includes demographics, diagnosis codes, laboratory measurements, bedside physiological observations, treatment records, and derived clinical variables. Data were queried from the MIMIC-IV tables in Google BigQuery and assembled into a patient-level analytic dataset for subsequent modeling in Python.

The prediction task was defined at a fixed landmark 24 hours after ICU admission. Clinical information recorded from ICU admission through this landmark was used to estimate subsequent in-hospital mortality among patients who remained alive and hospitalized at 24 hours. This design was chosen to align cohort eligibility, predictor availability, and outcome assessment to a common prediction time [23].

The analysis used a fixed development and locked-test design. The final cohort was divided before preprocessing or model fitting, and all feature preprocessing, feature selection, hyperparameter optimization, calibration, and operating-threshold selection were performed using the development data only. The locked test set was used for final performance assessment and did not feed back into these development procedures. The analytical and reporting framework was informed by current guidance for clinical prediction modeling [24].

MIMIC-IV is distributed through PhysioNet under its applicable data-use and credentialing requirements. The database contains de-identified records collected under the institutional approvals governing MIMIC-IV. This study involved secondary analysis of de-identified data and no direct interaction with patients. The overall analytical workflow, including cohort definition, development-only model construction, and locked test-set evaluation, is summarized in Figure 1.

**Figure 1.**
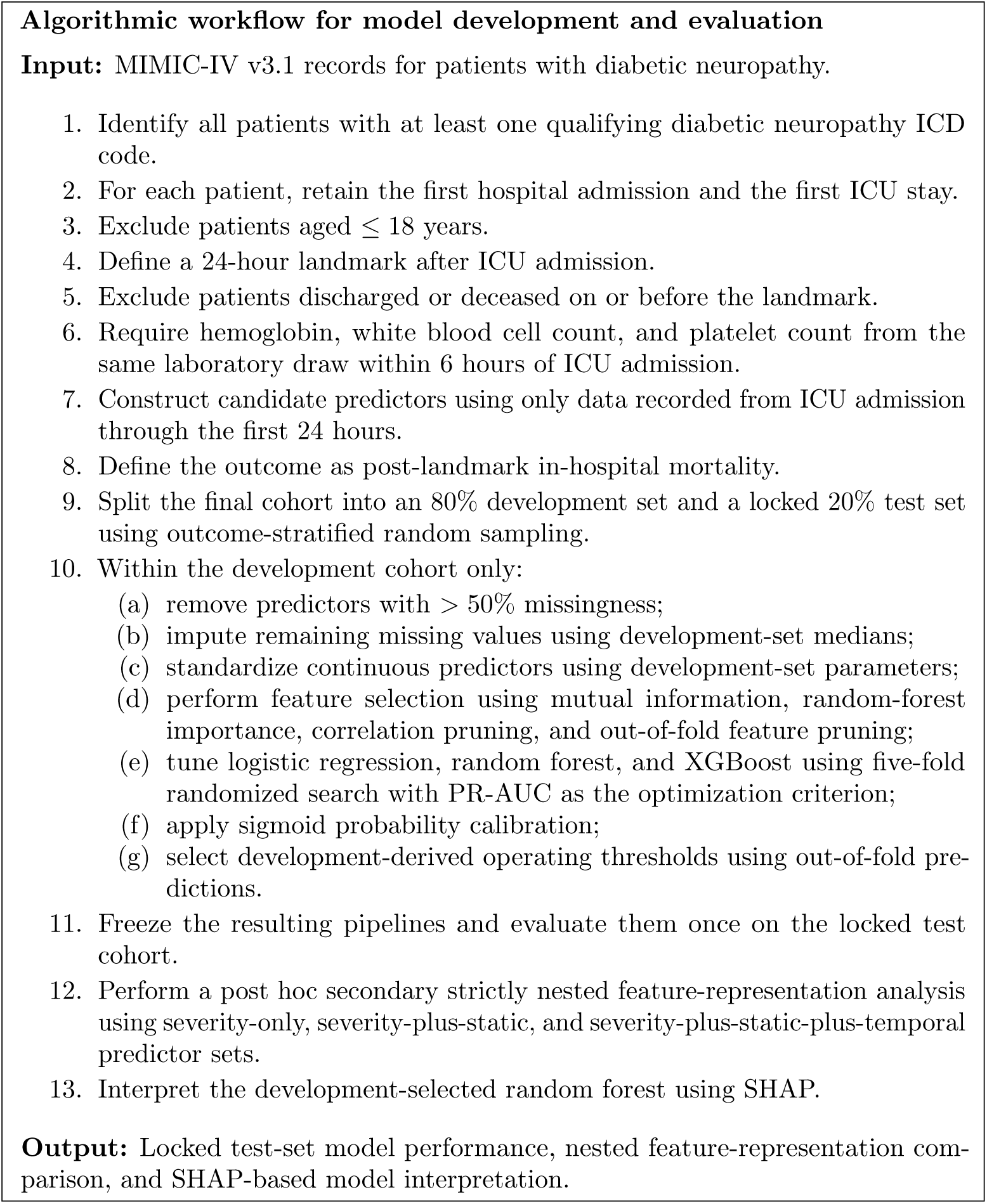
Pseudo-code summary of the analytical workflow. All preprocessing, feature selection, hyperparameter optimization, probability calibration, and threshold selection were restricted to the development cohort. The locked test cohort was not used for model-development decisions and was used for primary performance assessment and the post hoc nested comparison.

### 2.2. Study Population and Prediction Landmark

Patients with diabetic neuropathy were identified from hospital diagnosis records. A patient was considered eligible for the diabetic-neuropathy cohort if any recorded hospital admission contained a qualifying International Classification of Diseases (ICD) diagnosis code. ICD-9 codes included 249.60, 249.61, and 250.60–250.63; ICD-10 codes included E10.40, E10.42, E10.43, E11.40–E11.43, and E11.49.

After identification of patients with diabetic neuropathy, the first hospital admission was selected for each patient, followed by the first ICU stay within that hospitalization. Patients aged 18 years or younger were excluded. The prediction landmark was defined as exactly 24 hours after ICU admission. To belong to the population at risk at this time, a patient was required to remain alive and hospitalized beyond the landmark. Patients who died or were discharged on or before ICU admission plus 24 hours were therefore excluded.

Eligible patients were additionally required to have hemoglobin, white blood cell count, and platelet count recorded from the same laboratory draw within 6 hours after ICU admission. This requirement preserved a consistent early complete-blood-count measurement framework for the analytic cohort.

The study outcome was all-cause in-hospital mortality occurring after the 24-hour landmark. Mortality was obtained from the MIMIC-IV hospital mortality indicator. Because patients who died before or at the landmark were excluded from the at-risk population, the resulting outcome should be interpreted as post-landmark in-hospital mortality rather than mortality from the time of ICU admission.

This landmark criterion avoids using subsequent hospitalization duration to determine eligibility for a prediction made at 24 hours. This distinction prevents patients from being selected according to information that becomes known only after the intended prediction time.

### 2.3. Candidate Predictors

Candidate predictors were derived from information available no later than 24 hours after ICU admission. The initial feature space included demographics, comorbidity indicators, disease-severity scores, Glasgow Coma Scale measurements, vital signs, laboratory measurements, selected early ICU interventions, and derived inflammatory indices.

Comorbidity variables included myocardial infarction, congestive heart failure, chronic pulmonary disease, renal disease, hypertension, obesity, and the Charlson Comorbidity Index. Illness severity was represented by available standard ICU severity measures, including Acute Physiology Score III (APS III), Simplified Acute Physiology Score II, Logistic Organ Dysfunction System (LODS), and Oxford Acute Severity of Illness Score. APS III and LODS were also retained separately for the severity-score comparison [9, 10].

Physiological and laboratory variables included routinely recorded vital signs, glucose measurements, renal and electrolyte markers, coagulation tests, and complete blood count measurements. Predictor extraction was timestamp-restricted so that measurements recorded after the 24-hour landmark were not incorporated into model features.

### 2.4. Temporal Feature Construction

Repeated clinical measurements during the first ICU day were represented using both conventional summaries and temporal descriptors. Depending on the underlying variable and its source table, the extracted features included the first and last observed values, mean, minimum, maximum, within-window standard deviation, number of measurements, change between the first and last measurements, and an indicator that no measurement was available during the observation window.

For a repeatedly measured variable *x*, change over the first ICU day was defined as

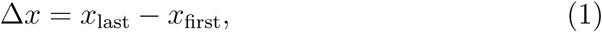

where *x*_first_ and *x*_last_ denote the first and final recorded values within the 24-hour observation window. Within-day variability was represented by the sample standard deviation of available observations,

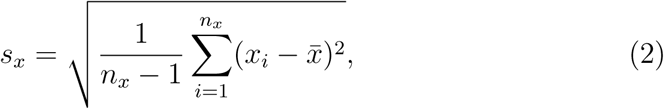

when more than one measurement was available. The measurement count *n_x_* was retained for selected variables because frequency of testing may carry additional information about clinical monitoring and disease severity.

The temporal representation was intended to preserve simple aspects of within-day evolution without requiring complete or regularly sampled time series. It therefore captures direction of change, variability, and measurement intensity while remaining compatible with conventional tabular prediction algorithms. These features should not be interpreted as direct measurements of disease progression, because both measurement timing and frequency may be influenced by clinical workflow.

### 2.5. Development and Locked Test Sets

The final cohort was divided into an 80% development set and a 20% locked test set using outcome-stratified random sampling with random seed 42. The development partition was used throughout preprocessing and model development. The test partition remained separate until the fitted pipelines and model-specific operating thresholds had been determined.

Predictors with more than 50% missingness in the development set were removed. For retained numeric predictors, missing values were imputed using medians estimated from the development data. Continuous predictors with more than two unique values were standardized using means and standard deviations estimated from the development set. The fitted imputation and scaling parameters were then applied unchanged to the locked test data. Sex was encoded as a binary predictor before model fitting.

This ordering ensured that missing-value replacement, scaling parameters, and feature availability were determined without using information from the locked test cohort.

### 2.6. Feature Selection

Feature selection was conducted exclusively within the development set. Two complementary ranking procedures were initially applied. Mutual information was calculated between each predictor and the mortality outcome, and predictors within the highest-ranking 25% were retained. In parallel, a class-weighted random forest with 100 trees was fitted to the development data, and the 40 predictors with the highest impurity-based feature importance were identified. The initial candidate set consisted of the intersection of these two groups. If the intersection was empty, the highest-ranking mutual-information features were retained as a fallback.

Redundant predictors were subsequently removed using pairwise absolute correlations. Candidate variables were considered in descending order of mutual information, and a variable was excluded when its absolute correlation with an already retained predictor exceeded 0.85.

When more than 15 predictors remained after this step, an additional leave-one-feature-out pruning procedure was performed using five-fold out-of-fold predictions within the development cohort. A class-weighted random forest was used as the pruning estimator. For each predictor, model AUROC after removing that predictor was compared with the full candidate model. Predictors whose removal reduced out-of-fold AUROC by no more than 0.002 were eligible for removal, with a maximum of eight predictors removed in this stage.

The resulting full feature representation contained 14 predictors and was fixed before evaluation on the locked test set.

### 2.7. Prediction Algorithms

Three prediction algorithms were evaluated: logistic regression, random forest, and XGBoost [25, 26]. Logistic regression provided a conventional linear benchmark, whereas random forest and XGBoost were included to represent nonlinear tree-based methods capable of modeling interactions among clinical variables.

Class imbalance was addressed during model fitting. Logistic regression and random forest used class-weighted learning, while XGBoost used a positive-class weight equal to the ratio of survivors to deaths in the development cohort.

Hyperparameters were tuned separately for each algorithm using randomized search with stratified five-fold cross-validation. Twelve randomly sampled parameter combinations were evaluated for each algorithm. For logistic regression, the search varied regularization strength, L2 or elastic-net penalty, and elastic-net mixing ratio. Random-forest tuning varied the number of trees, maximum tree depth, minimum leaf size, and number of candidate predictors considered at each split. XGBoost tuning varied tree depth, learning rate, number of boosting iterations, row subsampling, and column subsampling.

Average precision was used as the cross-validation optimization criterion. Throughout this study, reported PR-AUC values correspond to average precision calculated from the precision–recall curve. PR-AUC was included because post-landmark mortality was uncommon and therefore provided information complementary to AUROC in this imbalanced setting [27].

### 2.8. Probability Calibration and Operating-Threshold Selection

After hyperparameter optimization, each fitted algorithm underwent sigmoid probability calibration. Calibration was estimated using stratified five-fold cross-validation within the development cohort.

Model-specific operating thresholds were selected from calibrated out-of-fold predictions generated within the development data. Candidate thresholds from 0.05 through 0.95 in increments of 0.01 were examined. Among thresholds achieving sensitivity of at least 0.70 in the development out-of-fold predictions, the threshold with the highest F1 score was selected. If no candidate threshold reached the sensitivity target, the threshold with the highest F1 score overall was used.

The resulting thresholds were frozen before test-set evaluation. Consequently, a sensitivity of at least 0.70 was a development-stage threshold-selection criterion and was not assumed to remain satisfied in the locked test cohort.

### 2.9. Post Hoc Nested Feature-Representation Analysis

A post hoc secondary analysis was performed to examine the contribution of temporal information using strictly nested predictor sets. This analysis did not replace or modify the primary 14-feature modeling analysis. Instead, the three predictor representations were constructed by progressively adding information to a common baseline.

The severity-only representation contained APS III and LODS. The severity-plus-static representation added three non-temporal predictors from the original selected feature set: maximum chloride, minimum partial thromboplastin time, and maximum partial thromboplastin time. The full nested representation then added the remaining 11 temporal predictors from the original 14-feature set. The resulting groups therefore contained 2, 5, and 16 predictors, respectively. The 16-feature set was exactly the union of APS III and LODS with the 14 predictors selected in the primary analysis. Although the three predictors in the severity-plus-static representation were derived from measurements collected during the first 24 hours, they were treated as non-temporal summary features because they did not explicitly encode measurement order, within-day change, variability, or measurement frequency.

Each nested predictor set was evaluated independently using logistic regression, random forest, and XGBoost. Hyperparameter tuning, sigmoid calibration, and operating-threshold selection were repeated within the development cohort using the same five-fold cross-validation and out-of-fold procedures as in the primary analysis. The resulting pipelines were then evaluated in the same locked test cohort.

Because the predictor sets were strictly nested, changes in performance across the three representations were used to assess whether the addition of temporal information improved prediction beyond severity scores and non-temporal first-day summaries. This comparison was treated as a secondary analysis and did not alter the original development-selected 14-feature models.

### 2.10. Model Performance and Statistical Analysis

Performance was assessed in the locked test cohort using discrimination, probability accuracy, calibration, and threshold-dependent classification measures. Discrimination was summarized using AUROC and PR-AUC. Because the mortality prevalence was low, PR-AUC was interpreted alongside AUROC rather than relying on AUROC alone [27].

Probability-prediction error was summarized using the Brier score. Calibration was quantified using calibration slope and calibration intercept, estimated by fitting a logistic model of the observed outcome against the logit of predicted mortality probability [28, 29].

At the model-specific operating thresholds derived from development out-of-fold predictions, sensitivity, specificity, positive predictive value, negative predictive value, and F1 score were calculated. These measures describe performance at the selected operating point and were interpreted separately from probability-level model performance.

Ninety-five percent confidence intervals for AUROC were estimated using 1,000 nonparametric bootstrap resamples of the locked test cohort with random seed 42. Each bootstrap sample contained the same number of observations as the original test cohort and was sampled with replacement. Resamples containing only one outcome class were discarded. The 2.5th and 97.5th percentiles of the bootstrap AUROC distribution were reported as the confidence limits.

### 2.11. Model Interpretability

The algorithm used for model interpretation was selected using development-set performance rather than locked-test results. Among the three primary 14-feature models, random forest had the highest mean five-fold cross-validated PR-AUC during hyperparameter optimization (0.375), compared with 0.347 for logistic regression and 0.335 for XGBoost. Random forest was therefore selected for the interpretability analysis before consideration of locked-test performance.

Model interpretation was performed using SHapley Additive exPlana-tions (SHAP) [30]. The prediction pipeline used for locked-test probabilities consisted of the tuned random forest followed by five-fold sigmoid calibration. For SHAP analysis, the underlying tuned RandomForestClassifier obtained from the randomized hyperparameter search was refitted to the full development cohort before probability calibration and interpreted using shap.TreeExplainer.

SHAP values were calculated for all 1,077 patients in the development cohort after applying preprocessing parameters fitted on the development data. The locked test cohort was not used to derive global feature importance. Overall predictor importance was summarized using the mean absolute SHAP value across development observations, while a beeswarm plot was used to display both the magnitude and direction of individual feature contributions.

Because SHAP was applied to the underlying random forest rather than the subsequent sigmoid-calibration layer, the resulting values describe contributions to the uncalibrated model output rather than to the final calibrated probabilities. SHAP results were interpreted as descriptions of model behavior and predictive associations, not as evidence of causal effects.

### 2.12. Software and Reproducibility

Cohort construction and feature extraction were implemented using structured query language in Google BigQuery. Statistical preprocessing and model development were performed in Python 3.14.3. The primary modeling workflow used scikit-learn 1.8.0, XGBoost 3.2.0, pandas 2.3.3, and NumPy 2.4.2. Model interpretation was performed using SHAP 0.51.0, and figures were generated using matplotlib 3.10.8. Serialized fitted model objects were managed using joblib 1.5.3.

A fixed random seed of 42 was used for the development/test split, cross-validation procedures, randomized hyperparameter search, feature-selection estimators, and bootstrap resampling where supported. The same locked development/test split was retained for the primary analysis and the secondary nested feature-representation analysis.

## 3. Results

### 3.1. Study Cohort

A total of 8,992 patients had at least one qualifying diabetic neuropathy diagnosis code in MIMIC-IV. After restricting the analysis to the first ICU stay during the first hospital admission, 1,876 patients remained. Thirty-six patients were discharged or died on or before the 24-hour landmark and were excluded from the population at risk; 12 of these patients died before the landmark. Among the 1,840 patients alive and still hospitalized at 24 hours, a further 493 were excluded because a same-draw hemoglobin, white blood cell count, and platelet count was not available within 6 hours of ICU admission.

The final landmark cohort included 1,347 patients, of whom 83 (6.16%) died after the landmark during the index hospitalization. The development cohort contained 1,077 patients and 66 deaths, while the locked test cohort contained 270 patients and 17 deaths (Figure 2).

**Figure 2.**
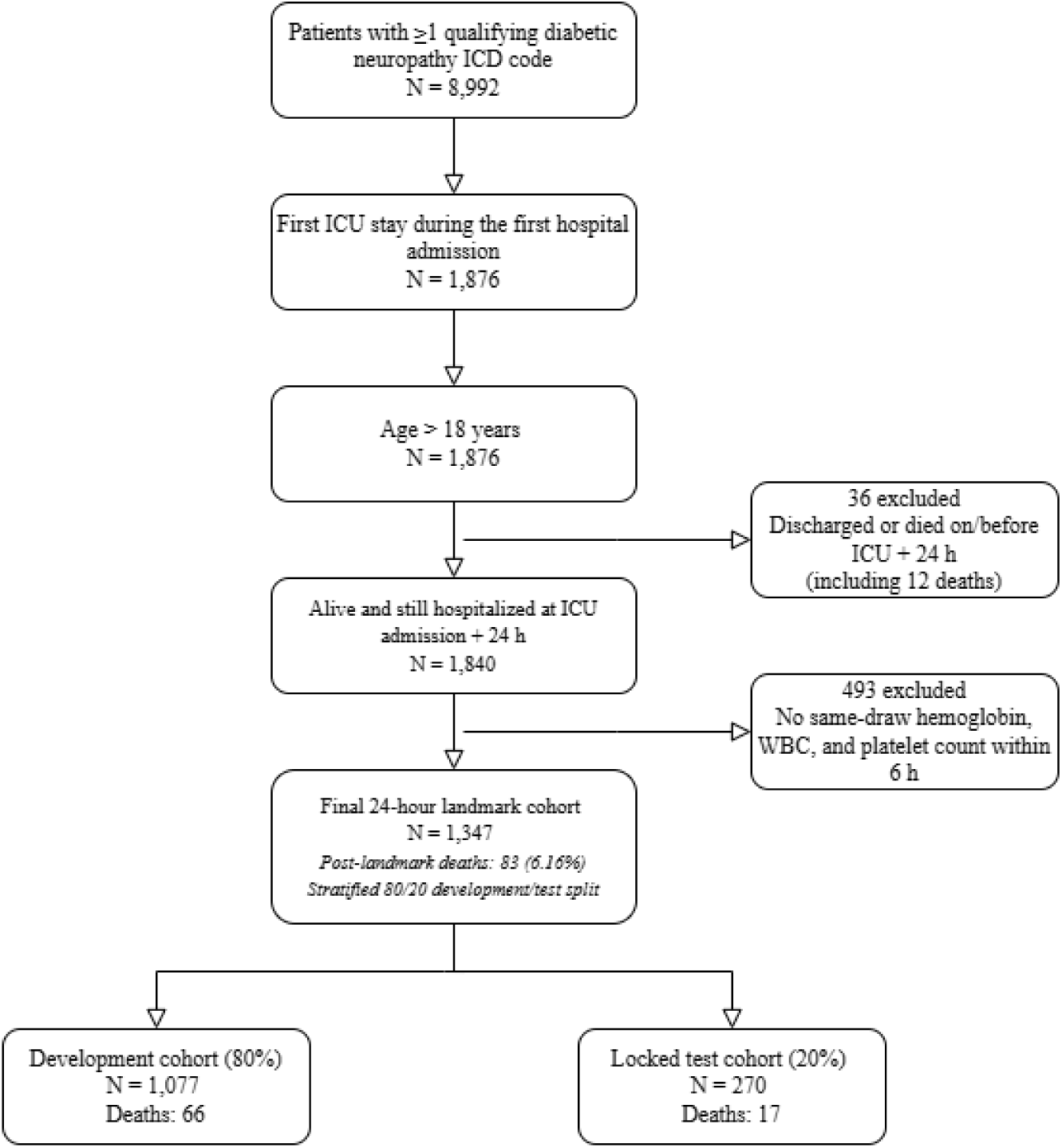
Study cohort selection and development–test allocation using the 24-hour landmark design. Patients discharged or deceased on or before ICU admission plus 24 hours were excluded from the population at risk for post-landmark in-hospital mortality. CBC, complete blood count; ICD, International Classification of Diseases; ICU, intensive care unit.

Baseline characteristics are summarized in Table 1. Non-survivors were older and had a greater burden of renal disease and overall comorbidity. Differences were most pronounced for illness-severity measures: mean APS III was 67.66 among non-survivors compared with 44.48 among survivors (SMD 1.004), while LODS was 7.37 versus 4.44 (SMD 0.926). Non-survivors also had higher first-day RDW, creatinine, and BUN values and were more likely to receive invasive ventilation and renal replacement therapy during the first 24 hours.

**Table 1.** Baseline characteristics of the final 24-hour landmark cohort, stratified by post-landmark in-hospital survival.

| Characteristic | Overall<br>(*n* = 1,347) | Survivors<br>(*n* = 1,264) | Non-survivors<br>(*n* = 83) | SMD |
| --- | --- | --- | --- | --- |
| *Demographics* |  |  |  |  |
| Age, years | 65.54 ± 12.84 | 65.34 ± 12.82 | 68.58 ± 12.79 | 0.253 |
| Male sex | 845 (62.7%) | 786 (62.2%) | 59 (71.1%) | 0.190 |
| *Comorbidities* |  |  |  |  |
| Myocardial infarction | 419 (31.1%) | 391 (30.9%) | 28 (33.7%) | 0.060 |
| Congestive heart failure | 459 (34.1%) | 424 (33.5%) | 35 (42.2%) | 0.179 |
| Chronic pulmonary disease | 310 (23.0%) | 289 (22.9%) | 21 (25.3%) | 0.057 |
| Renal disease | 488 (36.2%) | 442 (35.0%) | 46 (55.4%) | 0.420 |
| Hypertension | 558 (41.4%) | 535 (42.3%) | 23 (27.7%) | -0.310 |
| Obesity | 357 (26.5%) | 336 (26.6%) | 21 (25.3%) | -0.029 |
| *Severity scores* |  |  |  |  |
| Charlson comorbidity index | 6.28 ± 2.50 | 6.17 ± 2.48 | 7.93 ± 2.29 | 0.736 |
| APS III | 45.91 ± 19.36 | 44.48 ± 17.82 | 67.66 ± 27.38 | 1.004 |
| SAPS II | 37.10 ± 13.09 | 36.21 ± 12.23 | 50.67 ± 17.60 | 0.955 |
| LODS | 4.62 ± 2.74 | 4.44 ± 2.57 | 7.37 ± 3.66 | 0.926 |
| OASIS | 31.61 ± 8.46 | 31.13 ± 8.16 | 38.92 ± 9.59 | 0.875 |
| *Glasgow Coma Scale* |  |  |  |  |
| Total GCS | 13.70 ± 2.80 | 13.77 ± 2.72 | 12.72 ± 3.65 | -0.324 |
| *Vital signs (first-day mean)* |  |  |  |  |
| Heart rate, beats/min | 84.37 ± 14.14 | 84.18 ± 13.92 | 87.30 ± 16.98 | 0.201 |
| Mean arterial pressure, mmHg | 77.94 ± 10.68 | 78.09 ± 10.75 | 75.60 ± 9.21 | -0.249 |
| Respiratory rate, breaths/min | 18.70 ± 3.50 | 18.58 ± 3.45 | 20.50 ± 3.81 | 0.526 |
| Temperature, °C | 36.85 ± 0.51 | 36.86 ± 0.46 | 36.80 ± 0.97 | -0.079 |
| Charted glucose, mg/dL, median [IQR] | 155.96<br>[132.08–201.47] | 154.30<br>[131.60–198.65] | 183.50<br>[146.35–223.98] | 0.317 |
| *Laboratory measurements (first day)* |  |  |  |  |
| Hemoglobin, g/dL | 10.38 ± 1.91 | 10.39 ± 1.90 | 10.15 ± 2.00 | -0.125 |
| White blood cell count, ×10<sup>9</sup>/L | 13.23 ± 7.75 | 12.97 ± 6.88 | 17.19 ± 15.52 | 0.352 |
| Platelet count, ×10<sup>9</sup>/L | 201.18 ± 89.86 | 201.29 ± 89.03 | 199.53 ± 102.30 | -0.018 |
| RDW, % | 14.78 ± 2.01 | 14.67 ± 1.92 | 16.40 ± 2.55 | 0.763 |
| Creatinine (maximum), mg/dL | 1.88 ± 1.99 | 1.81 ± 1.96 | 2.92 ± 2.15 | 0.539 |
| BUN (maximum), mg/dL | 32.80 ± 25.67 | 31.45 ± 24.50 | 53.63 ± 33.35 | 0.758 |
| *Interventions (first 24 h)* |  |  |  |  |
| Invasive ventilation | 608 (45.1%) | 556 (44.0%) | 52 (62.7%) | 0.381 |
| Any ventilation | 1089 (80.8%) | 1013 (80.1%) | 76 (91.6%) | 0.332 |
| Any vasopressor | 502 (37.3%) | 466 (36.9%) | 36 (43.4%) | 0.133 |
| Any renal replacement therapy | 83 (6.2%) | 66 (5.2%) | 17 (20.5%) | 0.468 |

*Note:* Continuous variables are presented as mean ± standard deviation unless otherwise indicated; categorical variables are *n* (%). Signed standardized mean differences (SMDs) compare non-survivors with survivors; negative values indicate lower values or prevalence among non-survivors. Charted glucose is presented as median [IQR] because of a single implausible bedside-glucose value; the glucose SMD excludes values > 1000 mg/dL. APS III, Acute Physiology Score III; BUN, blood urea nitrogen; GCS, Glasgow Coma Scale; LODS, Logistic Organ Dysfunction System; OASIS, Oxford Acute Severity of Illness Score; RDW, red cell distribution width; SAPS II, Simplified Acute Physiology Score II.

### 3.2. Selected Predictors

Feature selection was performed using the development cohort only. Fourteen predictors were retained for the full model: last blood urea nitrogen; calcium measurement count and standard deviation; glucose standard deviation; maximum chloride; first creatinine; change in heart rate; first and last international normalized ratio; last mean blood pressure; last, minimum, and maximum partial thromboplastin time; and first red cell distribution width. The selected predictors were dominated by renal, electrolyte, coagulation, and hemodynamic measurements recorded during the first 24 hours. Several variables represented within-day variability or change rather than a single measurement, including calcium and glucose variability, measurement frequency, and change in heart rate.

Among the three algorithms trained with this 14-feature representation, random forest had the highest development cross-validated PR-AUC (0.375), compared with 0.347 for logistic regression and 0.335 for XGBoost. Random forest was therefore selected for subsequent model-interpretability analysis without reference to locked-test performance.

### 3.3. Locked Test-Set Performance

All three models were evaluated once on the locked test cohort of 270 patients. Random forest achieved the highest AUROC at 0.851 (95% CI 0.765–0.924), with a PR-AUC of 0.339 and a Brier score of 0.051. XGBoost showed similar discrimination, with an AUROC of 0.847 (95% CI 0.771–0.914), PR-AUC of 0.307, and Brier score of 0.052. Logistic regression had an AUROC of 0.806 (95% CI 0.708–0.892), PR-AUC of 0.231, and Brier score of 0.056.

At the thresholds selected from out-of-fold development predictions, random forest had a sensitivity of 0.824 and specificity of 0.783. XGBoost had a sensitivity of 0.765 and specificity of 0.747, while logistic regression had a sensitivity of 0.647 and specificity of 0.822. Positive predictive values were low for all three models, ranging from 0.169 to 0.203, consistent with the low mortality prevalence in the test cohort. Negative predictive values ranged from 0.972 to 0.985.

Calibration slopes were 1.114 for random forest, 1.162 for XGBoost, and 0.825 for logistic regression. The corresponding calibration intercepts were 0.244, 0.330, and *−*0.418, respectively. Full performance estimates are shown in Table 2. Receiver operating characteristic and precision–recall curves are shown in Figure 3, and test-set calibration is shown in Figure 4.

**Figure 3.**
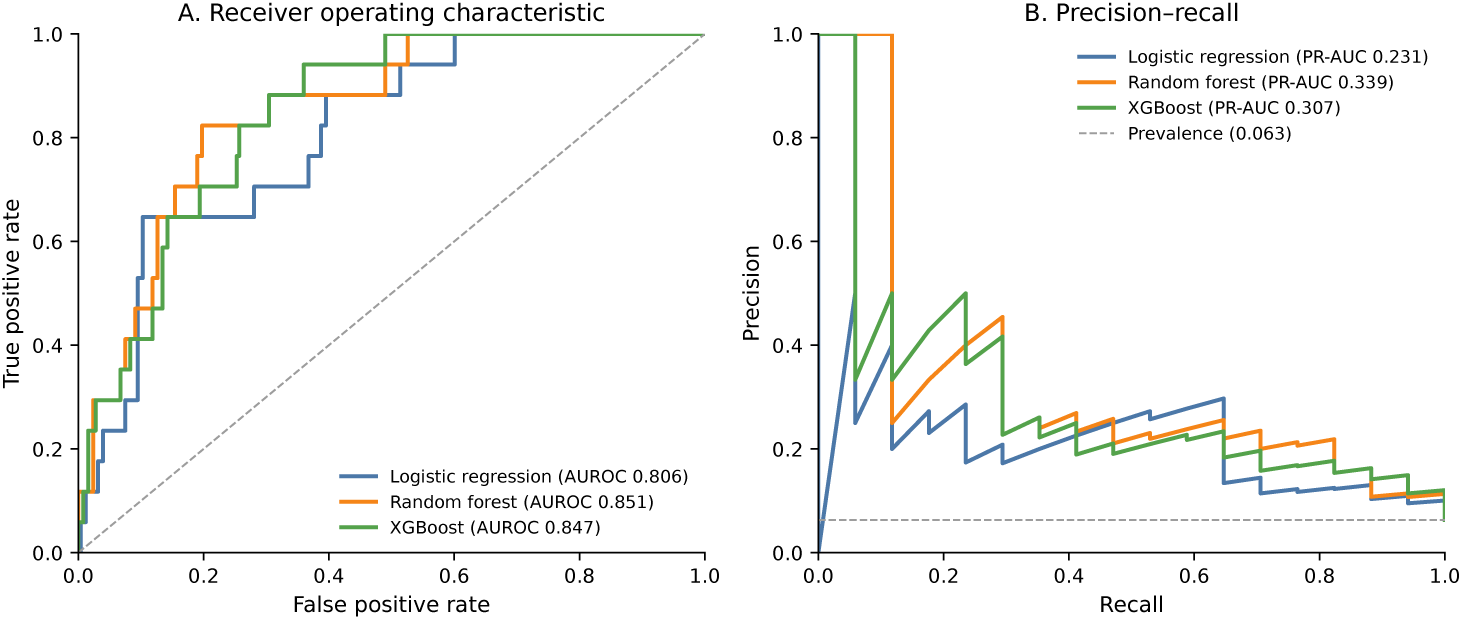
Discrimination of the three primary 14-feature models in the locked test cohort. (A) Receiver operating characteristic curves. (B) Precision–recall curves. All curves were generated using predictions from the locked test set (n = 270; 17 post-landmark deaths).

**Figure 4.**
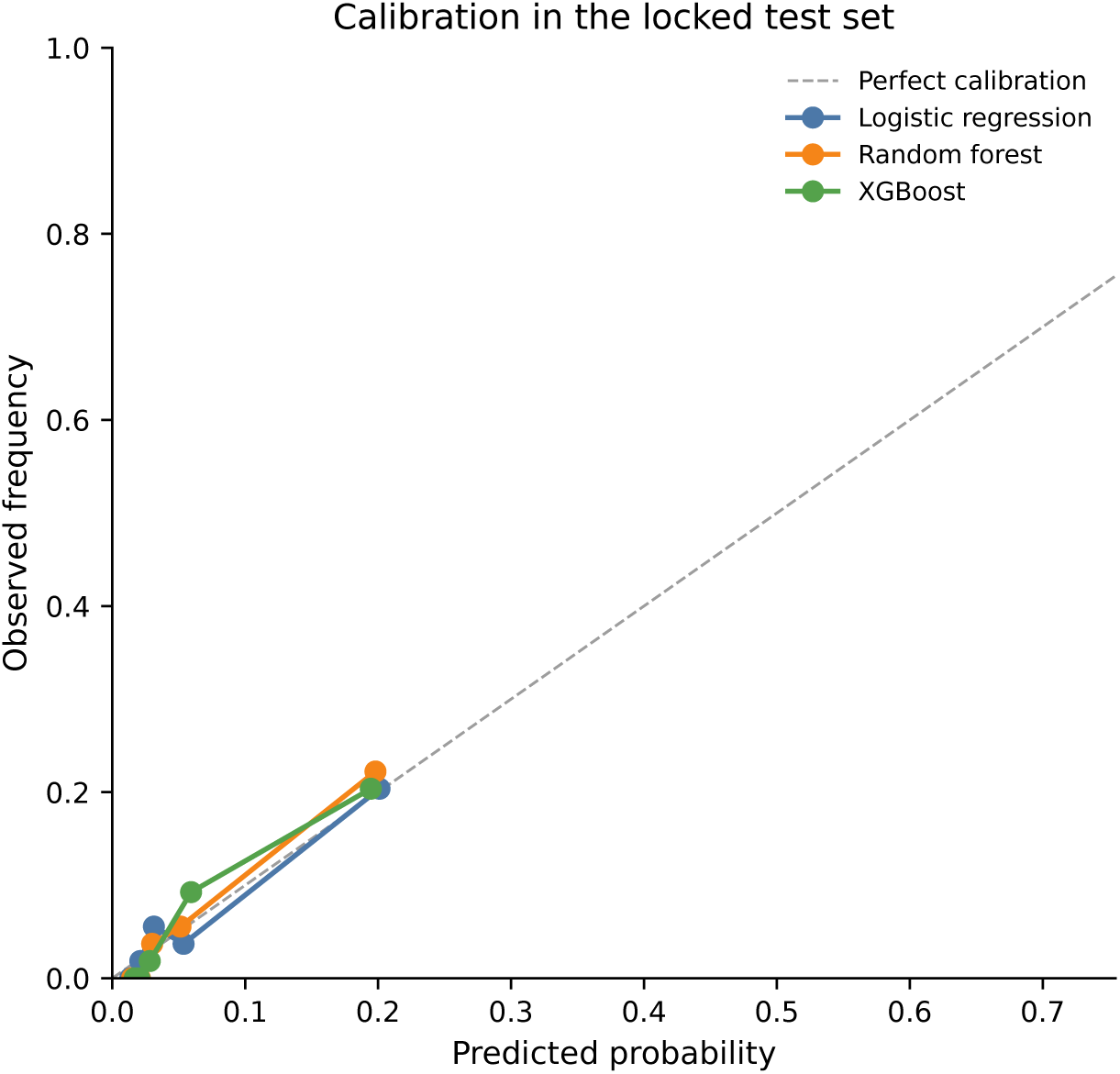
Calibration of predicted post-landmark in-hospital mortality risk in the locked test cohort. Observed event proportions are shown across quantile-based groups of predicted probability. The diagonal line represents perfect calibration.

**Table 2.** Performance of the three primary 14-feature models on the locked test set.

*Panel A. Discrimination and probability-level performance*
| Model | AUROC (95% CI) | PR-AUC | Brier score |
| --- | --- | --- | --- |
| Random forest | 0.851 (0.765–0.924) | 0.339 | 0.051 |
| XGBoost | 0.847 (0.771–0.914) | 0.307 | 0.052 |
| Logistic regression | 0.806 (0.708–0.892) | 0.231 | 0.056 |

*Panel B. Performance at the development-derived operating threshold*
| Model | Threshold | Sensitivity | Specificity | PPV | NPV | F1 |
| --- | --- | --- | --- | --- | --- | --- |
| Random forest | 0.06 | 0.824 | 0.783 | 0.203 | 0.985 | 0.326 |
| XGBoost | 0.06 | 0.765 | 0.747 | 0.169 | 0.979 | 0.277 |
| Logistic regression | 0.07 | 0.647 | 0.822 | 0.196 | 0.972 | 0.301 |

*Note:* Operating thresholds were selected using calibrated out-of-fold predictions from the development cohort and were frozen before evaluation on the locked test set. AUROC, area under the receiver operating characteristic curve; NPV, negative predictive value; PPV, positive predictive value; PR-AUC, area under the precision–recall curve.

### 3.4. Post Hoc Nested Feature-Representation Analysis

In the post hoc secondary nested analysis, discrimination increased as static and temporal information was progressively added to the severity-score baseline (Table 3; Figure 5). With APS III and LODS alone, logistic regression had the highest AUROC at 0.823 (95% CI 0.730–0.897). After adding the three static predictors, the highest observed AUROC was 0.833 (95% CI 0.748–0.909) with XGBoost.

**Figure 5.**
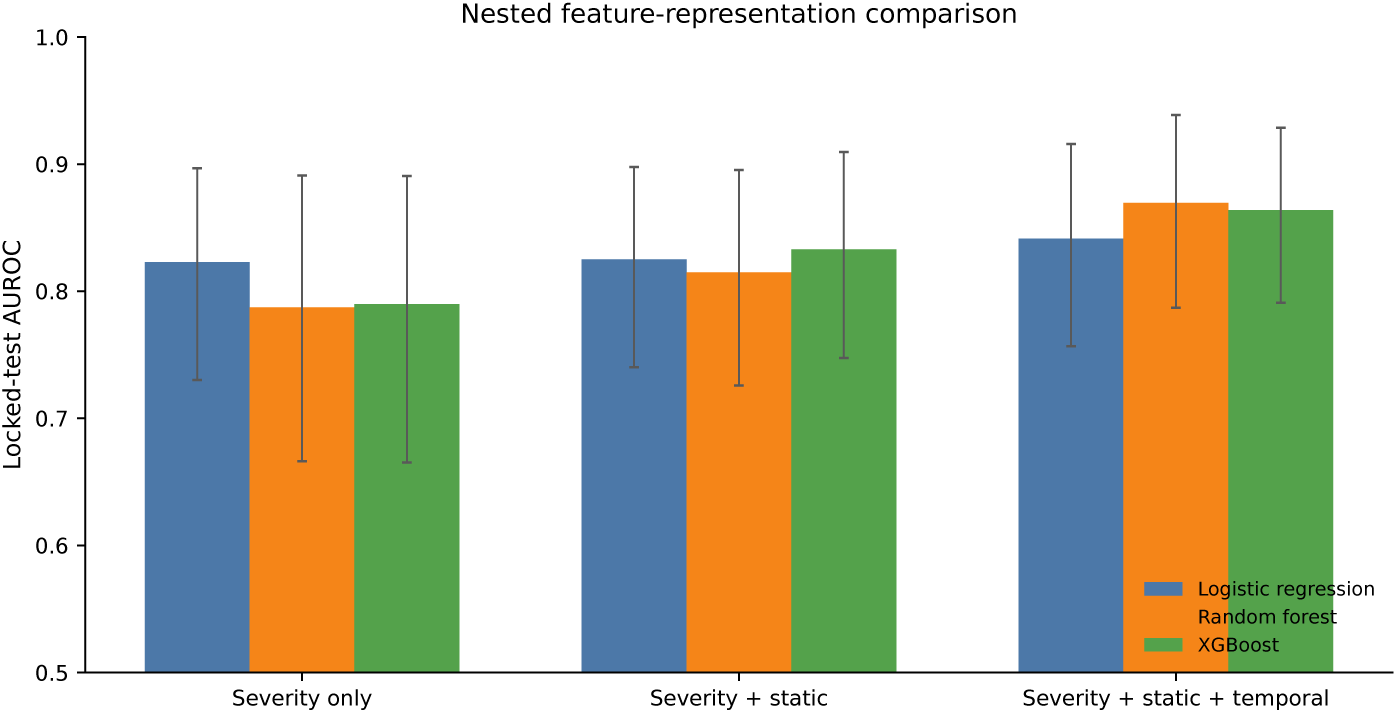
Post hoc comparison of locked test-set AUROC across strictly nested predictor representations. Error bars indicate 95% bootstrap confidence intervals. The severity-plus-static representation added three static predictors to APS III and LODS, while the full representation further added 11 temporal predictors.

**Table 3.** Locked test-set performance in the post hoc strictly nested predictor-representation analysis.

| Feature representation | Model | No. features | AUROC (95% CI) | PR-AUC | Brier |
| --- | --- | --- | --- | --- | --- |
| Severity only<br>(APS III + LODS) | Logistic regression | 2 | 0.823 (0.730–0.897) | 0.205 | 0.055 |
|  | Random forest | 2 | 0.787 (0.666–0.891) | 0.225 | 0.055 |
|  | XGBoost | 2 | 0.790 (0.665–0.891) | 0.211 | 0.055 |
| Severity + static | Logistic regression | 5 | 0.825 (0.740–0.898) | 0.245 | 0.055 |
|  | Random forest | 5 | 0.815 (0.726–0.895) | 0.333 | 0.053 |
|  | XGBoost | 5 | 0.833 (0.748–0.909) | 0.330 | 0.052 |
| Severity + static + temporal | Logistic regression | 16 | 0.841 (0.757–0.916) | 0.309 | 0.054 |
|  | Random forest | 16 | 0.870 (0.787–0.939) | 0.436 | 0.048 |
|  | XGBoost | 16 | 0.864 (0.791–0.929) | 0.407 | 0.051 |

*Note:* Predictor sets were strictly nested. The severity-only representation contained APS III and LODS; the static representation additionally contained maximum chloride and minimum and maximum partial thromboplastin time; the full representation further added the 11 temporal predictors from the primary selected feature set.

Adding the 11 temporal predictors produced the highest performance for all three algorithms relative to their corresponding severity-plus-static models. Logistic-regression AUROC increased from 0.825 to 0.841, random-forest AUROC from 0.815 to 0.870, and XGBoost AUROC from 0.833 to 0.864.

The same pattern was evident for PR-AUC, which increased from 0.245 to 0.309 for logistic regression, from 0.333 to 0.436 for random forest, and from 0.330 to 0.407 for XGBoost.

Among the fully nested models, random forest achieved the highest observed performance, with an AUROC of 0.870 (95% CI 0.787–0.939), PR-AUC of 0.436, and Brier score of 0.048. These results support a modest but consistent gain in predictive performance when temporal first-day information was added to severity scores and non-temporal summaries.

### 3.5. Model Interpretation

SHAP analysis was performed for the development-selected 14-feature random forest using the full development cohort. The number of calcium measurements during the first ICU day had the highest mean absolute SHAP value (0.061), followed by last BUN (0.059), first creatinine (0.039), first RDW (0.036), and last partial thromboplastin time (0.034). Other relatively influential predictors included minimum partial thromboplastin time, last INR, maximum partial thromboplastin time, within-day glucose variability, and last mean blood pressure.

The ranking included both absolute clinical measurements and temporal descriptors. In particular, measurement frequency, within-day variability, and first-to-last change were retained alongside renal and coagulation markers. The SHAP beeswarm plot further demonstrated heterogeneous and nonlinear contributions across individual patients (Figure 6). These findings describe associations learned by the fitted random forest and should not be interpreted as causal effects.

**Figure 6.**
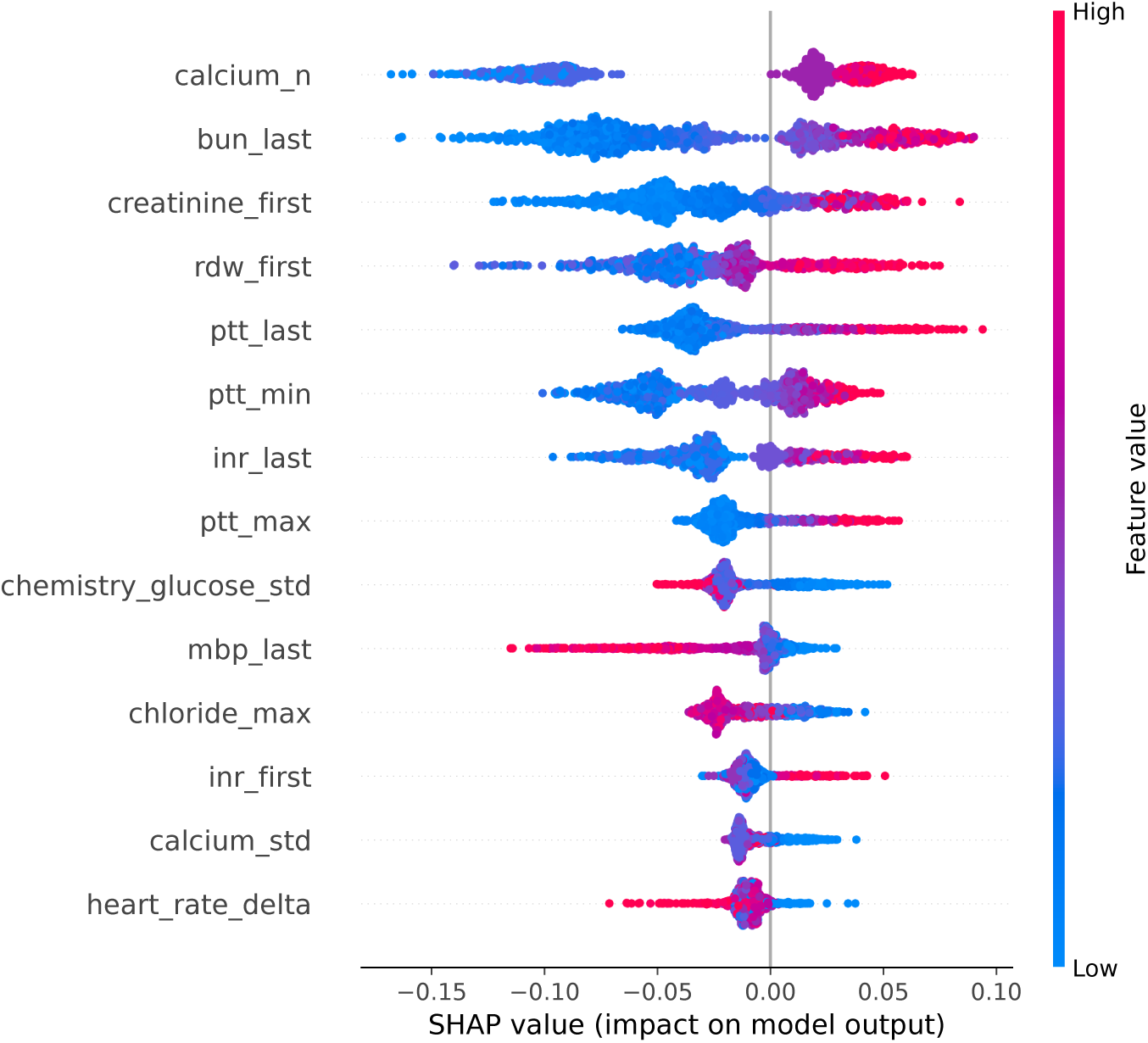
SHAP summary plot for the development-selected 14-feature random forest. Predictors are ordered by mean absolute SHAP value across the development cohort (n = 1,077). Each point represents one patient-feature contribution; color indicates the observed feature value, and horizontal position indicates the contribution to the uncalibrated random-forest output.

## 4. Discussion

### 4.1. Principal Findings

In this retrospective MIMIC-IV study, we evaluated post-landmark in-hospital mortality among ICU patients with diabetic neuropathy using information collected during the first 24 hours of ICU care. The analysis was anchored to a 24-hour landmark so that cohort eligibility, predictor collection, and outcome assessment referred to the same prediction time. Model development was also separated from final evaluation: preprocessing, feature selection, hyperparameter tuning, calibration, and operating-threshold selection were completed using the development cohort before the locked test set was evaluated.

Among the primary 14-feature models, random forest had the highest development cross-validated PR-AUC and was therefore selected for model interpretation without reference to locked-test performance. On the locked test cohort, random forest achieved an AUROC of 0.851, PR-AUC of 0.339, and Brier score of 0.051. XGBoost showed similar discrimination, with an AUROC of 0.847, whereas logistic regression achieved an AUROC of 0.806. The overlapping confidence intervals do not support a clear performance difference between the two tree-based algorithms.

The post hoc nested analysis provided a complementary assessment of the value of temporal information. When predictors were added progressively to the same baseline representation, discrimination improved after temporal variables were introduced for each of the three algorithms. Random-forest AU-ROC increased from 0.815 using severity scores and non-temporal summary predictors to 0.870 using the full temporal representation, while PR-AUC increased from 0.333 to 0.436. Corresponding AUROC increases were observed for logistic regression (0.825 to 0.841) and XGBoost (0.833 to 0.864). These differences were modest in absolute terms, but their direction was consistent across algorithms. The results therefore support the presence of additional prognostic information in first-day temporal measurements beyond that contained in severity scores and the selected non-temporal summary variables.

### 4.2. Comparison With Previous Studies

Longitudinal clinical data have increasingly been used for risk prediction in critical care. Prior studies have shown that repeated physiological and laboratory measurements can support dynamic outcome prediction and preserve information that may be lost when an ICU admission is represented using only baseline or aggregate measurements [11, 12, 13]. Related studies in patients with diabetes have also linked early glycemic trajectories and glucose variability with subsequent clinical outcomes [15, 16]. The present results are consistent with this broader literature, while focusing on a narrower population of ICU patients with diabetic neuropathy.

Huang et al. previously developed machine-learning models for mortality prediction among ICU patients with diabetic neuropathy using MIMIC-IV and reported an AUROC of approximately 0.78 for their best-performing random forest model [22]. Their work established the feasibility of mortality prediction in this population. The present study addresses a different aspect of the prediction problem by explicitly defining a 24-hour prediction landmark, separating development from locked evaluation, and comparing nested representations of severity, non-temporal summaries, and temporal first-day information.

Numerical performance should not be compared directly across the two studies. Differences in cohort construction, prediction timing, predictor definitions, feature engineering, and evaluation procedures can materially alter AUROC and other performance estimates. The main distinction of the present analysis is therefore methodological rather than a claim that one algorithm is intrinsically superior. Our findings indicate that, when the prediction task is aligned to a 24-hour landmark, simple summaries of within-day evolution can contribute information beyond non-temporal first-day summaries.

### 4.3. Clinical and Methodological Interpretation

The predictors retained by the primary random-forest model were concentrated in renal, coagulation, electrolyte, and hemodynamic measurements. SHAP analysis identified the number of calcium measurements, last blood urea nitrogen, first creatinine, first red cell distribution width, and last partial thromboplastin time among the most influential features. Both absolute measurements and temporal descriptors appeared among the higher-ranking predictors.

These findings should be interpreted cautiously. For example, the strong contribution of calcium measurement frequency does not imply that repeated calcium testing itself changes mortality risk. Measurement frequency can reflect clinical concern, treatment intensity, evolving organ dysfunction, or local practice patterns. Similarly, renal and coagulation variables may act as markers of overall physiological instability rather than disease-specific mechanisms. SHAP values describe how the fitted model uses the observed data; they do not establish causal relationships between individual predictors and mortality.

The nested analysis is useful in this respect because it evaluates temporal representation at the level of predictive performance rather than relying on the importance assigned to any single temporal feature. Adding the temporal predictors improved both AUROC and PR-AUC across logistic regression, random forest, and XGBoost. This consistency suggests that the additional information was not dependent on one particular modeling algorithm. At the same time, the magnitude of improvement was not large enough to suggest that temporal summaries replace established measures of illness severity. A more reasonable interpretation is that they complement severity scores and non-temporal summaries.

Threshold-dependent results also illustrate an important limitation of discrimination metrics alone. Post-landmark mortality occurred in only 6.16% of the cohort. At the development-derived operating threshold, the primary random forest achieved sensitivity of 0.824 and specificity of 0.783, but its positive predictive value was only 0.203. Negative predictive value was 0.985. This pattern is expected when the target outcome is uncommon and reinforces the need to consider PR-AUC, calibration, and operating characteristics alongside AUROC [27, 28]. The present model is therefore better viewed as a risk-stratification model than as a stand-alone trigger for treatment decisions.

### 4.4. Strengths

A major strength of this study is the explicit alignment of the prediction task to a clinically interpretable time point. Patients were required to remain alive and hospitalized at 24 hours, predictors were restricted to information available through that landmark, and mortality was assessed subsequently. This avoids defining eligibility using information that becomes available only later in the hospitalization and is consistent with the logic of landmark prediction [23].

The modeling workflow also maintained separation between development and final evaluation. Missing-data handling, scaling, feature selection, hyperparameter optimization, probability calibration, and operating-threshold selection were performed using the development cohort. The locked test set was reserved for final performance assessment. Random forest was selected for interpretation using development cross-validated PR-AUC rather than its subsequent locked-test AUROC. This reduces the risk that model choice or interpretation was influenced by repeated inspection of test-set results and is consistent with current prediction-model reporting guidance [24].

### 4.5. Limitations

This study has several limitations. First, all patients were drawn from MIMIC-IV, which represents care at a single academic medical center. The locked test set provides internal validation of the specified modeling workflow but does not establish transportability to other hospitals. Differences in case mix, laboratory testing, documentation, and treatment patterns may affect performance elsewhere.

We explored the feasibility of external evaluation using eICU-CRD but did not consider the available data sufficiently harmonized for formal external validation. Comparable diabetic-neuropathy phenotyping was sparse, and several predictors and severity measures could not be reproduced using definitions equivalent to those in MIMIC-IV. We therefore chose not to present those results as evidence of external validation. Independent validation remains necessary.

Second, the number of outcome events was limited. The final cohort contained 83 post-landmark deaths, including only 17 in the locked test cohort. Confidence intervals were therefore relatively wide, and threshold-dependent estimates such as sensitivity and positive predictive value remain imprecise. The nested comparison also demonstrates consistent directional improvement rather than a formal statistical test of pairwise differences in AUROC or PR-AUC.

Third, diabetic neuropathy was defined using diagnosis codes recorded in the electronic health record. Coding may miss clinically recognized neuropathy and does not provide consistent information on neuropathy subtype, duration, or severity. The findings therefore apply to a coded diabetic-neuropathy cohort rather than a prospectively adjudicated phenotype.

Fourth, 493 otherwise eligible patients were excluded because a same-draw hemoglobin, white blood cell count, and platelet count was not available within 6 hours after ICU admission. This requirement ensured consistent availability of the early complete-blood-count framework used in cohort construction, but it may have selected patients who underwent more intensive early testing.

Fifth, temporal information was represented using summary features such as first and last values, variability, measurement count, and first-to-last change. These descriptors are simple and reproducible but cannot capture the full structure of irregularly sampled ICU time series. More complex longitudinal models may extract additional information, although any increase in complexity would need to produce reproducible gains over simpler baselines.

Sixth, SHAP analysis was applied to the tuned random forest before sigmoid probability calibration and was calculated in the development cohort. It therefore describes feature contributions to the underlying random-forest output rather than directly to calibrated probabilities. In addition, SHAP explanations remain model-dependent associations and should not be interpreted causally.

Finally, this was a retrospective prediction study. Good discrimination or calibration in historical data does not demonstrate that displaying model predictions will improve clinical decisions or patient outcomes. Clinical usefulness would need to be evaluated prospectively. The nested feature-representation analysis was specified after completion of the primary analysis and reused the same locked test cohort; these secondary comparisons should therefore be interpreted as exploratory.

### 4.6. Future Work

The next step is external validation in a cohort where both the diabetic-neuropathy phenotype and the required first-day predictors can be reproduced consistently. Such evaluation should preserve the 24-hour landmark and apply the fitted model without using the external outcomes for model redevelopment. Discrimination and calibration should both be examined, with local recalibration considered separately if required.

Future work could also evaluate richer longitudinal representations, including irregular time-series models or repeated landmark predictions that update risk as additional ICU data become available. These approaches should be compared directly with the simpler temporal summaries used here rather than assuming that increased model complexity improves clinically useful prediction.

Prospective evaluation would ultimately be required to determine whether 24-hour mortality-risk estimates add useful information to clinical assessment and whether acting on those estimates improves care. Such studies should define how predictions would change monitoring or treatment before assessing clinical impact.

## 5. Conclusion

Using a 24-hour landmark design and a locked internal test cohort, we developed machine-learning models for subsequent in-hospital mortality among ICU patients with diabetic neuropathy. The development-selected 14-feature random forest achieved an AUROC of 0.851 on the locked test set, with discrimination similar to XGBoost and higher than logistic regression in this cohort.

In a post hoc strictly nested analysis, adding temporal first-day predictors to severity scores and non-temporal summaries improved discrimination across all three evaluated algorithms. The largest observed performance was obtained with the full nested random-forest model, which achieved an AUROC of 0.870 and PR-AUC of 0.436. These findings suggest that simple representations of within-day clinical evolution can provide prognostic information beyond non-temporal first-day summaries.

These findings are based on internal validation within a single data source. External validation, assessment of transportability across institutions, and prospective evaluation of clinical utility are required before these models can be considered for clinical use.

## CRediT authorship contribution statement

Janet Sanjaya: Data curation, Formal analysis, Methodology, Software, Validation, Visualization, Writing – original draft, Writing – review and editing. Sakshie Pathak: Conceptualization, Methodology, Writing – review and editing. Yong Si: Methodology, Validation, Visualization, Writing – review and editing. Mohammadsaeed Haghi: Conceptualization, Methodology, Writing – review and editing. Nausin Tabassum Kudrot: Conceptualization, Methodology, Writing – review and editing. Greg Placencia: Writing – review and editing. Kamiar Alaei: Writing – review and editing. Maryam Pishgar: Conceptualization, Project administration, Supervision, Writing – review and editing. All authors reviewed and approved the final manuscript.

## Ethics approval and consent to participate

This study used de-identified data from MIMIC-IV version 3.1. Access was obtained through PhysioNet after completion of the required credentialing, human-subjects research training, and applicable data-use agreements. Because the study involved secondary analysis of de-identified records and no direct interaction with participants, no additional institutional review board approval or individual informed consent was required.

## Data availability

The MIMIC-IV version 3.1 data analyzed in this study are available to credentialed researchers through PhysioNet, subject to completion of the required research training and data-use agreements. Because these are third-party controlled-access data, patient-level records and derived patient-level extracts cannot be redistributed by the authors. No new patient data were collected.

## Code availability

The analysis code supporting the findings of this study may be obtained from the corresponding author upon reasonable request, subject to the applicable PhysioNet data-use requirements.

## Acknowledgments

The authors have no additional acknowledgments.

## Declaration of generative AI and AI-assisted technologies in the manuscript preparation process

During the preparation of this work, the authors used OpenAI ChatGPT for editorial restructuring, language refinement, and drafting assistance, and Anthropic Claude for language refinement. After using these tools, the authors reviewed and edited the content as needed, verified all numerical and methodological statements against the original analyses and source materials, and take full responsibility for the content of the publication. The tools were not used to generate or modify patient data, construct study cohorts, perform statistical analyses, train or evaluate the machine learning models, or generate the reported results.

## References

[1] R. Pop-Busui, A. J. M. Boulton, E. L. Feldman, V. Bril, R. Freeman, R. A. Malik, J. M. Sosenko, D. Ziegler, Diabetic neuropathy: A position statement by the american diabetes association, Diabetes Care 40 (1) (2017) 136–154. doi:10.2337/dc16-2042.

[2] E. L. Feldman, B. C. Callaghan, R. Pop-Busui, D. W. Zochodne, D. E. Wright, D. L. Bennett, V. Bril, J. W. Russell, V. Viswanathan, Diabetic neuropathy, Nature Reviews Disease Primers 5 (2019) 41.doi:10.1038/s41572-019-0092-1.

[3] S. Tesfaye, A. J. M. Boulton, P. J. Dyck, R. Freeman, M. Horowitz, P. Kempler, G. Lauria, R. A. Malik, V. Spallone, A. Vinik, L. Bernardi, P. Valensi, Toronto Diabetic Neuropathy Expert Group, Diabetic neuropathies: update on definitions, diagnostic criteria, estimation of severity, and treatments, Diabetes Care 33 (10) (2010) 2285–2293.doi:10.2337/dc10-1303

[4] R. Pop-Busui, L. Ang, A. J. M. Boulton, E. L. Feldman, R. L. Marcus, K. Mizokami-Stout, J. R. Singleton, D. Ziegler, Diagnosis and Treatment of Painful Diabetic Peripheral Neuropathy, ADA Clinical Compendia Series, American Diabetes Association, Arlington, VA, 2022. doi:10.2337/db2022-01

[5] D. Selvarajah, D. Kar, K. Khunti, M. J. Davies, A. R. Scott, J. Walker, S. Tesfaye, Diabetic peripheral neuropathy: advances in diagnosis and strategies for screening and early intervention, The Lancet Diabetes & Endocrinology 7 (12) (2019) 938–948.doi:10.1016/S2213-8587(19)30081-6

[6] American Diabetes Association Professional Practice Committee for Diabetes, 12. retinopathy, neuropathy, and foot care: Standards of care in diabetes—2026, Diabetes Care 49 (Supplement 1) (2026) S261–S276.doi:10.2337/dc26-S012

[7] W.-C. Hsu, S. Y.-H. Chiu, A. M.-F. Yen, L.-S. Chen, C.-Y. Fann, C.-S. Liao, H.-H. Chen, Somatic neuropathy is an independent predictor of all-and diabetes-related mortality in type 2 diabetic patients: a population-based 5-year follow-up study (kcis no. 29), European Journal of Neurology 19 (9) (2012) 1192–1198. doi:10.1111/j.1468-1331.2011.03659.x.

[8] J.-L. Vincent, R. Moreno, Clinical review: Scoring systems in the critically ill, Critical Care 14 (2) (2010) 207. doi:10.1186/cc8204.

[9] W. A. Knaus, D. P. Wagner, E. A. Draper, J. E. Zimmerman, M. Bergner, P. G. Bastos, C. A. Sirio, D. J. Murphy, T. Lotring, A. Damiano, et al., The APACHE III prognostic system: risk prediction of hospital mortality for critically ill hospitalized adults, Chest 100 (6) (1991) 1619–1636.doi:10.1378/chest.100.6.1619

[10] J.-R. Le Gall, J. Klar, S. Lemeshow, F. Saulnier, C. Alberti, A. Artigas, D. Teres, ICU Scoring Group, The logistic organ dysfunction system: a new way to assess organ dysfunction in the intensive care unit, JAMA 276 (10) (1996) 802–810. doi:10.1001/jama.1996.03540100046027.

[11] H. Harutyunyan, H. Khachatrian, D. C. Kale, G. Ver Steeg, A. Galstyan, Multitask learning and benchmarking with clinical time series data, Scientific Data 6 (2019) 96. doi:10.1038/s41597-019-0103-9.

[12] H.-C. Thorsen-Meyer, A. B. Nielsen, A. P. Nielsen, B. S. Kaas-Hansen, P. Toft, J. Schierbeck, T. Strøm, A. Chmura, M. Heimann, L. Dybdahl, M. Spangsege, P. B. Cordua, O. Winther, Dynamic and explainable machine learning prediction of mortality in patients in the intensive care unit: a retrospective study of high-frequency data in electronic patient records, The Lancet Digital Health 2 (4) (2020) e179–e191.doi:10.1016/S2589-7500(20)30018-2

[13] S. L. Hyland, M. Faltys, M. Hüser, X. Lyu, T. Gumbsch, C. Esteban, C. Bock, M. Horn, M. Moor, B. Rieck, M. Zimmermann, D. Bodenham, K. Borgwardt, G. Rätsch, T. M. Merz, et al., Early prediction of circulatory failure in the intensive care unit using machine learning, Nature Medicine 26 (2020) 364–373. doi:10.1038/s41591-020-0789-4.

[14] Y. Si, L. Sun, S. Chen, J. Fan, E. Pishgar, K. Alaei, G. Placencia, M. Pishgar, Retrospective machine learning approach for forecasting in-hospital death in icu patients after cardiac arrest, Informatics in Medicine Unlocked 64 (2026) 101776. doi:10.1016/j.imu.2026.101776.

[15] H. Ye, Z. Ma, Q. Zong, Q. Zhu, Y. Yan, S. Yang, P. Xiang, H. Zou, Association of the time in targeted blood glucose range of 140 to 180 mg/dl with the mortality of critically ill patients with diabetes: analysis of the MIMIC-IV database, Journal of Clinical & Translational Endocrinology 41 (2025) 100413. doi:10.1016/j.jcte.2025.100413.doi:10.1016/j.jcte.2025.100413.

[16] D. Kong, Y. Long, T. Huang, Y. Zhang, W.-J. Liu, C.-X. Liu, S. Liu, Early ICU glycemic trajectory phenotypes predict short- and long-term mortality in critically ill patients: a dual-database cohort study, Metabolism Open 30 (2026) 100478. doi:10.1016/j.metop.2026.100478.

[17] A. Rajkomar, E. Oren, K. Chen, A. M. Dai, N. Hajaj, M. Hardt, P. J. Liu, X. Liu, J. Marcus, M. Sun, P. Sundberg, H. Yee, K. Zhang, Y. Zhang, G. Flores, G. E. Duggan, J. Irvine, Q. Le, K. Litsch, et al., Scalable and accurate deep learning with electronic health records, npj Digital Medicine 1 (2018) 18. doi:10.1038/s41746-018-0029-1.

[18] B. Shickel, P. J. Tighe, A. Bihorac, P. Rashidi, Deep ehr: A survey of recent advances in deep learning techniques for electronic health record (ehr) analysis, IEEE Journal of Biomedical and Health Informatics 22 (5) (2018) 1589–1604. doi:10.1109/JBHI.2017.2767063.

[19] A. E. W. Johnson, L. Bulgarelli, L. Shen, A. Gayles, A. Shammout, S. Horng, T. J. Pollard, S. Hao, B. Moody, B. Gow, L.- w. H. Lehman, L. A. Celi, R. G. Mark, MIMIC-IV, a freely accessible electronic health record dataset, Scientific Data 10 (2023) 1.doi:10.1038/s41597-022-01899-x

[20] A. E. W. Johnson, L. Bulgarelli, L. Shen, A. Gayles, A. Shammout, S. Horng, T. J. Pollard, B. Moody, B. Gow, L.-w. H. Lehman, L. A. Celi, R. G. Mark, MIMIC-IV (version 3.1), 10.13026/kpb9-mt58. URL PhysioNet (2024). doi: 10.13026/kpb9-mt58 URL 10.13026/kpb9-mt58

[21] S. Chen, Y. Si, J. Fan, L. Sun, G. Placencia, E. Pishgar, K. Alaei, M. Pishgar, Interpretable machine learning model for early prediction of 30-day mortality in icu patients with coexisting hypertension and atrial fibrillation: A retrospective cohort study, Informatics in Medicine Unlocked 59 (2025) 101717. doi:10.1016/j.imu.2025.101717. doi:10.1016/j.imu.2025.101717.

[22] Y. Huang, W. Mo, Y. Liu, S. Wang, Y. Zeng, P. Jiang, H. Wang, J. Bi, Machine learning models predict mortality risk in diabetic neuropathy patients using MIMIC-IV data, Scientific Reports 15 (2025) 38702. doi:.10.1038/s41598-025-22363-x

[23] H. C. van Houwelingen, Dynamic prediction by landmarking in event history analysis, Scandinavian Journal of Statistics 34 (1) (2007) 70–85.doi:10.1111/j.1467-9469.2006.00529.x

[24] G. S. Collins, K. G. M. Moons, P. Dhiman, R. D. Riley, A. L. Beam, B. Van Calster, M. Ghassemi, X. Liu, J. B. Reitsma, et al., TRIPOD+AI statement: updated guidance for reporting clinical prediction models that use regression or machine learning methods, BMJ 385 (2024) e078378.doi:10.1136/bmj-2023-078378

[25] L. Breiman, Random forests, Machine Learning 45 (1) (2001) 5–32. doi:10.1023/A:1010933404324

[26] T. Chen, C. Guestrin, XGBoost: A scalable tree boosting system, in: Proceedings of the 22nd ACM SIGKDD International Conference on Knowledge Discovery and Data Mining, ACM, New York, NY, USA, 2016, pp. 785–794. doi:10.1145/2939672.2939785.

[27] T. Saito, M. Rehmsmeier, The precision-recall plot is more informative than the ROC plot when evaluating binary classifiers on imbalanced datasets, PLOS ONE 10 (3) (2015) e0118432. doi:10.1371/journal.pone.0118432.

[28] E. W. Steyerberg, A. J. Vickers, N. R. Cook, T. Gerds, M. Gonen, N. Obu-chowski, M. J. Pencina, M. W. Kattan, Assessing the performance of prediction models: a framework for traditional and novel measures, Epidemiology 21 (1) (2010) 128–138. doi:10.1097/EDE.0b013e3181c30fb2.

[29] B. Van Calster, D. J. McLernon, M. van Smeden, L. Wynants, E. W. Steyerberg, Topic Group ‘Evaluating diagnostic tests and prediction models’ of the STRATOS initiative, Calibration: the achilles heel of predictive analytics, BMC Medicine 17 (1) (2019) 230.doi:10.1186/s12916-019-1466-7.

[30] S. M. Lundberg, S.-I. Lee, A unified approach to interpreting model predictions, in: Advances in Neural Information Processing Systems, Vol. 30, 2017, pp. 4765–4774. URL https://proceedings.neurips.cc/paper/2017/hash/8a20a8621978632d76c43dfd28b67767-Abstract.html

